# A phenome-wide multi-directional Mendelian randomization analysis of atrial fibrillation

**DOI:** 10.1101/2020.10.15.20212654

**Authors:** Qin Wang, Tom G Richardson, Eleanor Sanderson, Mika Ala-Korpela, George Davey Smith, Michael V Holmes

**Author notes:** Corresponding authors: Dr Qin Wang, Prof Michael V Holmes, Nuffield Department of Population Health, Big Data Institute Building, University of Oxford, Old Road Campus, Roosevelt Drive, Oxford OX3 7LF.

## Abstract

**Background:** The prevalence of atrial fibrillation (AF) is increasing with an aging worldwide population, yet a comprehensive understanding of its causes and consequences remains limited.

**Objectives:** To assess the causes and consequences of AF via a multi-directional Mendelian randomization (MR) analysis scanning thousands of traits in a hypothesis-free approach.

**Methods:** We used publicly available GWAS data centralised and harmonised by the IEU open GWAS database. We assessed the potential causal role of 5048 exposures on risk of AF and the causal role of genetic liability to AF on 10,308 outcomes via two-sample MR analysis. Multivariable MR analysis was further conducted to explore the comparative role of identified risk factors.

**Results:** MR analysis suggested that 55 out of 5048 exposure traits, including four proteins, play a causal role in AF (P < 1e-5 allowing for multiple comparisons). Multivariable analysis suggested that higher body mass index, height, systolic blood pressure as well as genetic liability to coronary artery diseases independently cause AF. Three out of the four proteins (DUSP13, TNFSF12 and IL6R) had a drug prioritising score for atrial fibrillation of 0.26, 0.38 and 0.88, respectively (values closer to 1 indicating stronger evidence of the protein as a potential drug target). Genetic liability to AF was linked to a higher risk of cardioembolic ischemic stroke.

**Conclusions:** Body mass index, height, systolic blood pressure and genetic liability to coronary artery diseases are independent causal risk factors for AF. Several proteins including DUSP13, IL-6R and TNFSF12 may represent therapeutic potential for preventing AF.

## Introduction

Atrial fibrillation (AF) is the most common cardiac rhythm disorder, affecting 1-2% of the population in Europe and North America.^1,2^ The prevalence and incidence of the AF is expected to increase further due to the aging population and it has been predicted that Europeans aged > 40 have a one in four lifetime risk of developing AF.^1^ AF is associated with an increased risk of stroke, myocardial infarction, heart failure, dementia and mortality, posing considerable challenges to public health and the economy.^1,3,4^

Despite remarkable advances in antiarrhythmic drugs, ablation procedures, and stroke-prevention strategies, AF remains an important cause of death and disability in middle-aged and elderly individuals.^5^ Clinical management of patients with AF is currently guided by stroke risk parameters, AF pattern, and symptoms.^5^ However, more than half of patients with AF remain symptomatic despite adequate anti-coagulation and rate control.^5^ Better understanding of the mechanisms leading to AF and the interplay of AF and its associated complications are warranted.

Observational studies have identified numerous risk factors to associate with AF risk, including obesity, smoking, alcohol consumption, diabetes, hypertension, reduced lung function, coronary artery diseases and heart failure.^1,4^ Mendelian randomization (MR) analyses^6^ have identified causal risk factors for AF, including higher fat mass,^7^ higher fat-free mass,^7^ higher birth weight,^8^ being taller^9^ and lower circulating soluble IL-6 receptors.^10^ On the other hand, studies assessing the consequences of genetic liability to AF are limited and a recent MR study reported a lack of a causal role of AF in Alzheimer’s.^11^ Despite these and other efforts geared at identifying individual risk factors for AF, studies applying a hypothesis-free approach to systemically identify the causes and consequences of AF have yet to be conducted. Here, we leveraged thousands of publicly available GWAS summary data and undertook a phenome-wide multi-directional MR analysis to comprehensively examine the causes and consequences of AF, which might provide an important basis to guide future strategies in preventing and treating AF, and avoiding AF-related sequelae.

## Methods

### Data sources

We used publicly available GWAS summary data, which are curated and centralised by MRC Integrative Epidemiology Unit (IEU) open GWAS database (https://gwas.mrcieu.ac.uk), and can be accessed via R package ‘TwoSampleMR’.^12,13^

### Traits filtering

There were 31,773 traits with GWAS summary data accessible via ‘TwoSampleMR’ package on date 18/04/2020. Traits pre-filtering was applied (**Figure 1A)** as follows: studies were included if they were primarily based on European descendants, had sample sizes over 3000 (to include as many traits as possible, e.g. to incorporate a proteomics GWAS (≈ 3000 proteins) with a sample size of 3301, and also have adequate sample sizes to generate reliable instruments),^13,14^ and had over 1 million genetic variants (to maximise the availability of the genetic instruments). In total, GWAS summary data for 3298 traits analysed using UK Biobank data released by Neale’s lab (second round) and MRC-IEU, and 7010 traits analysed by other consortiums or studies, are include here. This filtering resulted in 5048 exposure and 10308 outcome traits for the MR analyses (**Figure 1A)**.

**Figure 1.**
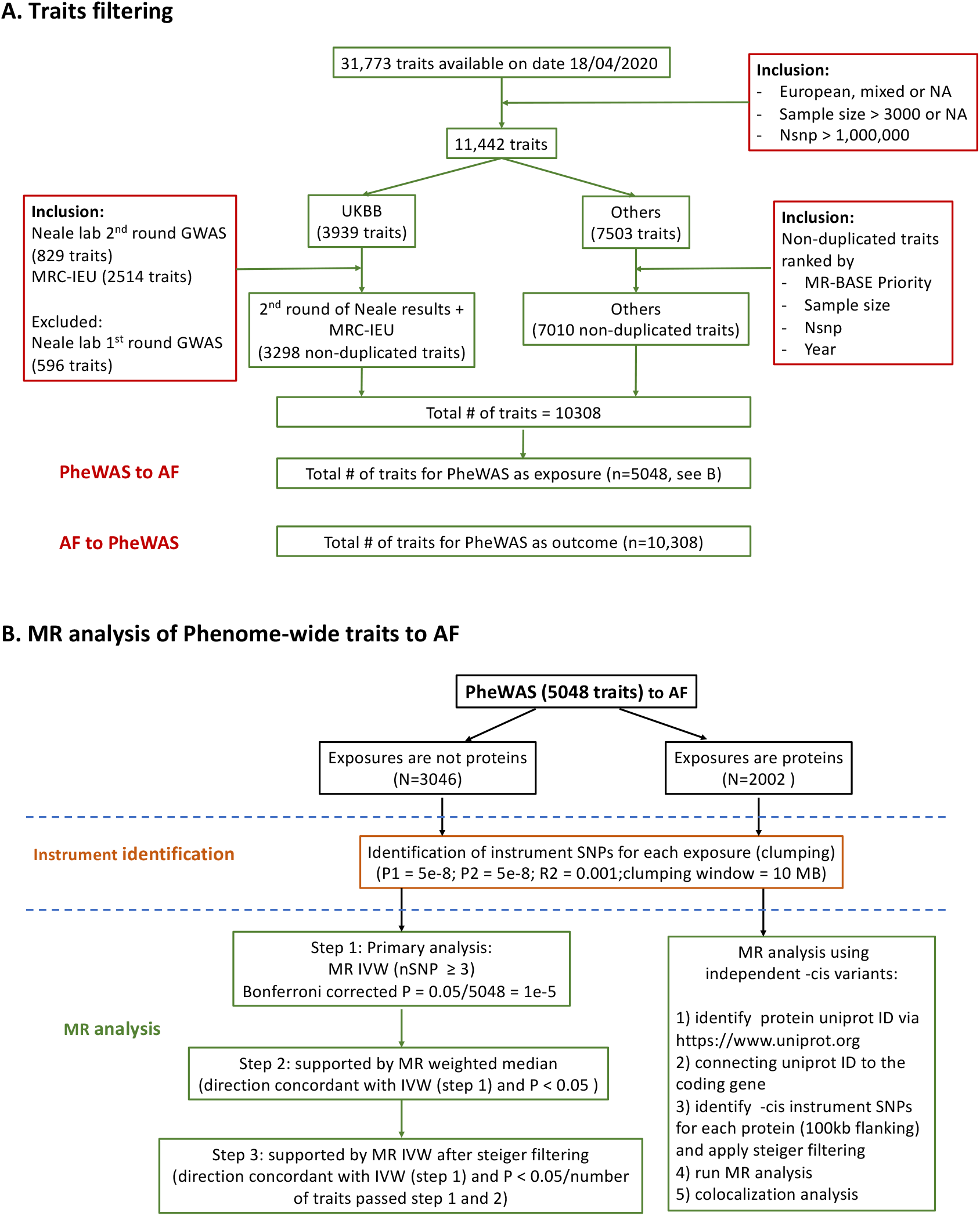
Analysis flow. A) traits filtering; B) analysis flow in conducting MR analysis of phenome-wide traits to atrial fibrillation.

### MR analysis from Phenome-wide traits (5048 exposures) to AF

The MR analysis flow of phenome-wide exposures to atrial fibrillation is shown in **Figure 1B**.

#### Genetic instruments for exposures

Clumping was applied to establish independent genetic variants for each individual exposure. Clumps are formed around central “index variants” which must have p-value no larger than 5e-8. Index variants were chosen greedily starting with the lowest p-value. Secondary hits were identified if they were within the clumping window (10Mb) of an index SNP, reached GWAS significance (P < 5e-8) and had a low LD with the index SNP (r2 < 0.001 based on 1000 Genomes phase 3 data from European descendants).

As binary traits from UK Biobank data were analysed in linear regression models, associations of the genetic variants with binary traits were scaled to log odds by multiplying a scaling factor 1/(μ*(1-μ)), where μ = n_case_/(n_case_ + n_conrol_).^14^ Whenever applicable, genetic associations with quantitative traits were reported in standard deviation (SD) and binary traits in log odds.

#### MR analysis

After genetic instruments were identified for each exposure, the associations of these genetic variants with atrial fibrillation were extracted. If the genetic variants were not directly available in the outcome GWAS, proxies with r2 > 0.8 were used based on 1000 Genomes phase 3 data from European descendants. In total, there were 5048 exposure-outcome pairs with instrument variants available in both exposure and outcome GWAS. Two sample MR analyses were performed via five different methods, including inverse-variance weighted (IVW), weighted median, MR Egger, simple mode and weighted mode. In general, genetic risk scores including multiple variants spanning the genome is preferred as the instrument for complex traits (e.g. non-protein measures), whist on the other hand *cis* variants located around the protein coding gene are typically considered as being more reliable instruments for proteins.^15^ Here, different MR analysis pipelines (**Figure 1B**) were used for protein vs. non-protein exposures to apply context-specific analytical approaches.

When exposures were complex traits (e.g. non-proteins), we selected all SNPs across the genome that associated with the trait at GWAS significance. MR estimates from the IVW method were treated as the primary results. The estimates were considered robust if they were supported by a three-stage approach: step 1 – there were more than three genetic variants for use in the instrument (minimal number of variants required to perform all five MR methods). Given the number of tests conducted, we used P < 0.05/ 5048 (number of total exposures) = 1e-5 as a heuristic to guide findings that were further explored; step 2 – primary IVW estimates were directionally concordant with those of weighted median, and P (weighted median) < 0.05; step 3 – primary IVW estimates were directionally concordant with IVW estimates after steiger filtering, and P (steiger) < 0.05 / number of traits passing steps 1 & 2. Steiger filtering was applied to ensure that each instrument variant explaining larger variances of exposures than outcomes, thus increasing the reliability of the assumed direction of causality. ^13^ For those exposure-outcome pairs that passed the three-stage sensitivity test, we further examined the consistency of the five MR methods.

When exposures were proteins, we aimed to identify -*cis* variants to proxy the proteins.^16^ We manually matched each protein to an unique Uniprot ID (https://www.uniprot.org). Then, we connected Uniprot ID to coding genes. For each protein, -*cis* instrument variants were identified if they were located within the coding gene (100kb flanking), were associated with the protein at P < 5e-8 and explained more variance in the proteins than the outcome trait. Here, we used Wald ratio or IVW estimates as the primary results. Similarly, Bonferroni corrected P < 0.05/5048 = 1e-5 was used to guide interpretation of the findings. For each protein, which was suggested to have a causal role for AF in the MR analysis, we further conducted colocalization analysis between the protein GWAS and AF GWAS at the protein coding gene (100kb flanking of the leading *cis*-pQTL) and used posterior probabilities of sharing one common causal variant (H4) to guide interpretation of colocalization.

### MR analysis from AF to Phenome-wide traits (10308 outcomes)

#### Genetic instruments for exposure (atrial fibrillation)

Using a similar approach as above, we identified 111 independent SNPs (between SNP LD r2< 0.001; association with AF P<5×10^−8^) as the genetic instruments for AF. GWAS summary data for AF is accessible via MRC-IEU database with ID = ebi−a−GCST006414.^17^

#### MR analysis

We extracted the associations of these 111 SNPs with each individual outcome trait. In total, there were 10308 exposure-outcome pairs with instrument variants available in both exposure and outcome GWAS. Similar to the above section described for non-protein exposures, we used IVW estimates as the primary results and results were considered robust only when they fulfilled the 3-stage analysis approach described above.

### Multivariable analyses

As many risk factors are typically correlated with each other, multivariable MR (MVMR) was used to explore independent causal risk factors to AF. We first grouped risk factors shown to have causal relationships with AF in univariable MR into different categories based on whether they were sharing the same feature (e.g. body mass index (BMI) and height were grouped in the category of anthropometry, whilst systolic and diastolic pressures (SBP, DBP) were grouped in the category of blood pressure). MVMR analyses were then conducted for traits within each individual category to elucidate their comparative causal role with risk of AF. The identified independent causal risk factors within each individual category were then selected and combined together in a final MVMR model to determine a set of credible independent causal risk factors for AF comprising traits across all categories. We estimated conditional F-statistics for the exposures in the MVMR models using the method described by Sanderson et al,^18^ and also provided the corresponding F-statistics in the UVMR models for a comparison.^19^

## Results

### MR analysis from phenome-wide traits to atrial fibrillation

Out of 5048 exposures (3046 non-protein measures and 2002 proteins), 55 traits were suggested as causal (Bonferroni P<1×10^−5^ from IVW MR) to the development of atrial fibrillation (**Figure 2-3**). The majority (50 of 55) were non-proteins, with most being related to anthropometry (e.g. height, fat mass, lean fat mass, waist, hip, height, birth weight, ankle spacing width, and impedance of leg). In general, positive relationships were identified between these anthropometric traits and AF risk, except impedance of legs. Fat-free mass (in arms, legs, trunk and whole body) displayed an approximately 1.5 times stronger magnitude (logOR per SD higher anthropometric trait) of relative risk than corresponding fat mass measures. In addition to anthropometric traits, higher basal metabolic rate, genetic liability to coronary artery disease (CAD), respiratory traits (forced expiratory volume in 1−second (FEV1) and forced vital capacity) and higher diastolic blood pressure were also linked to higher AF risk. In addition, a moderate causal effect was found for systolic blood pressure (P = 1.2e-5). In sensitivity analyses, the identified causal risk markers were consistently associated with AF across the six different MR methods (five MR methods using all genetic variants and one using steiger filtered variants (IVW)) (**Figure S1)**. The causal estimates for all 5048 exposures across the six methods are reported in **Table S1**. In addition, our results suggested that conventional cardiovascular risk factors including apolipoprotein B, triglycerides, LDL cholesterol, glucose, HbA1C and type 2 diabetes were not causal for AF (**Table S2**).

**Figure 2.**
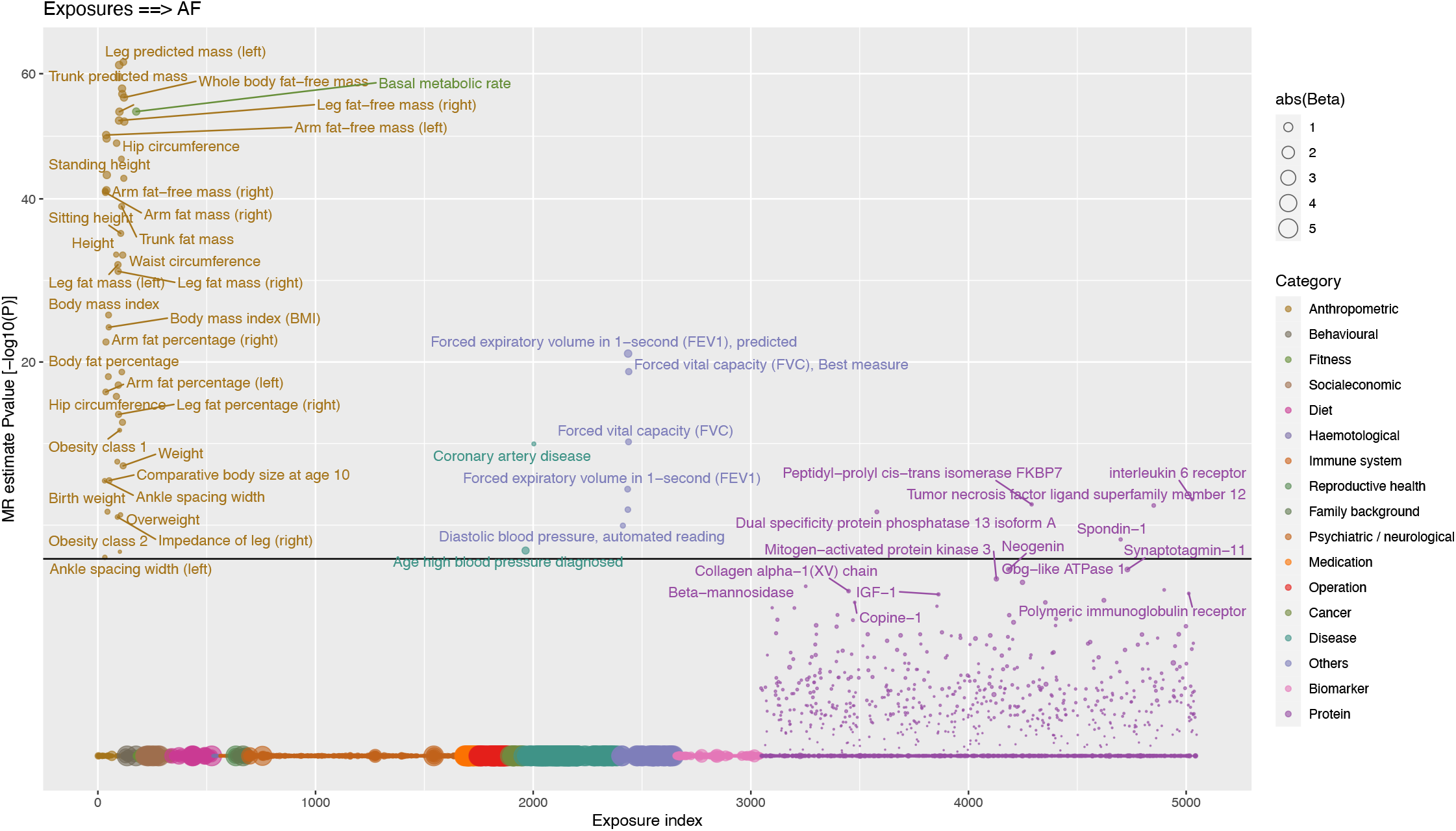
MR analysis of phenome-wide traits to atrial fibrillation. Y axis denotes the common logarithm (log10) of the P-values of the MR estimates and X axis denotes the number or index of the exposure traits. Symbols are coloured according the category of exposure traits and the symbol sizes are proportional to the absolute values of the MR estimates (based on IVW method). Horizontal black line corresponds to the Bonferroni corrected P = 0.05/5048 traits = 1e-5. For display purposes, the associations which were not supported by any of the three-stage test, we set the P values equal to one (indicating a lack of reliability).

**Figure 3.**
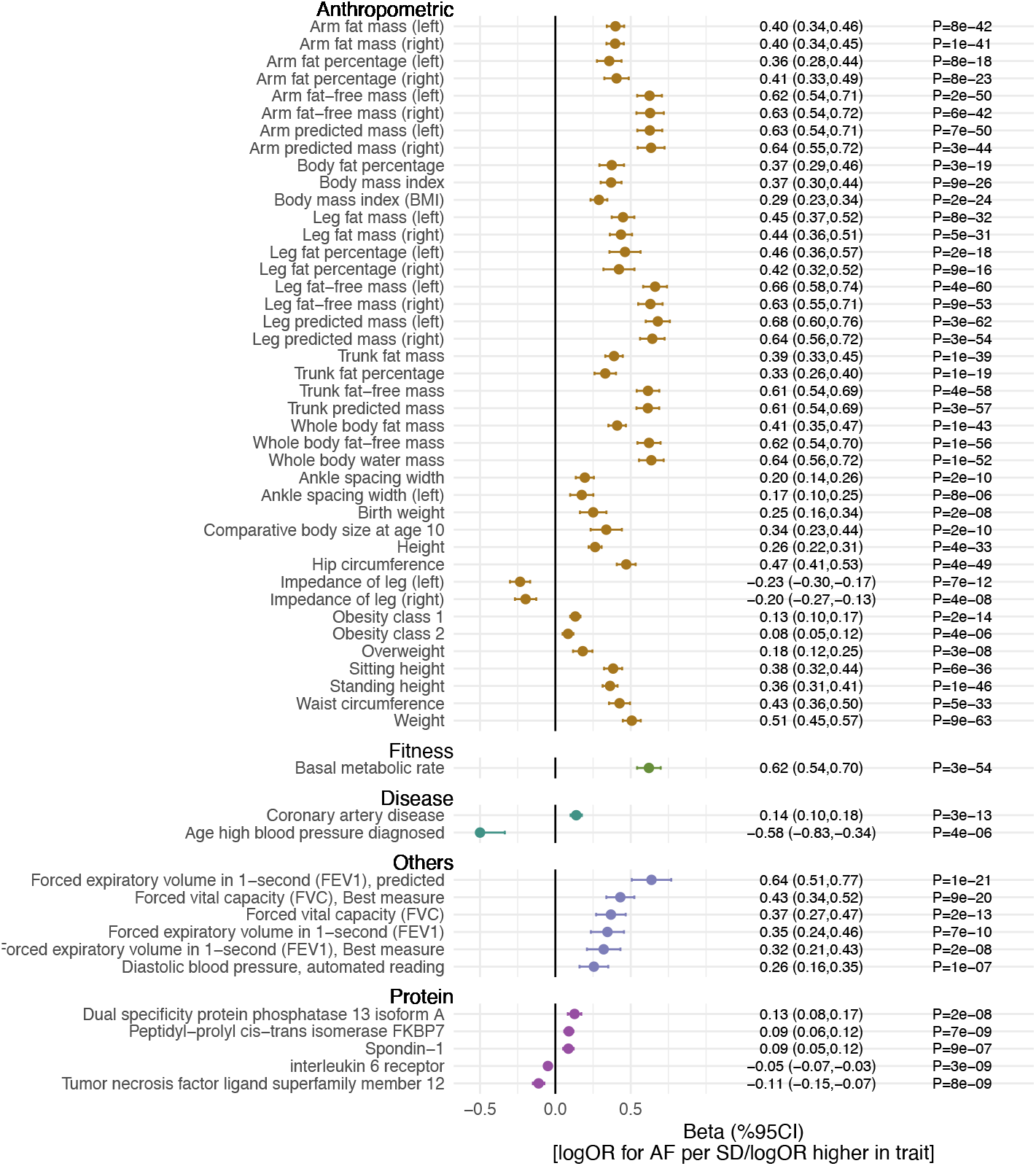
Causal risk factors for atrial fibrillation. The associations are reported as differences in log(OR) of AF per unit higher in the exposure trait. Traits presented are those that surpass multiple testing (Bonferroni P< 0.05/ 5048 exposures = 1e-5) in Figure 2.

Multivariable MR was used to investigate the comparative causal effects of the identified causal risk factors (**Figure 4**). We first fitted a multivariable model including markers showing evidence of causation with AF on univariable MR that were related to anthropometry and the results implicated BMI, hip and height showing independent causal relationships (**Figure 4A**). Similarly, we fitted a multivariable model for blood pressure traits and the results suggested that SBP was the underlying causal risk factor (**Figure 4B**). Finally, we selected the above identified casual markers (BMI, hip circumferences, height and systolic blood pressure) and generated a multivariable model which also included other markers, including ankle spacing width, impedance of leg, basal metabolic rate, birth weight, CAD and forced expiratory volume (**Figure 4C**), and the model identified BMI, height, SBP and liability to CAD to be the traits displaying direct causal effects on risk of AF (**Figure 4C-D**). The genetic correlations of these causal risk factors are reported in **Figure S2** and the conditional F-statistics of these exposures across the MVMR models and corresponding F-statistics in the UVMR models are listed in **Table S3**.

**Figure 4.**
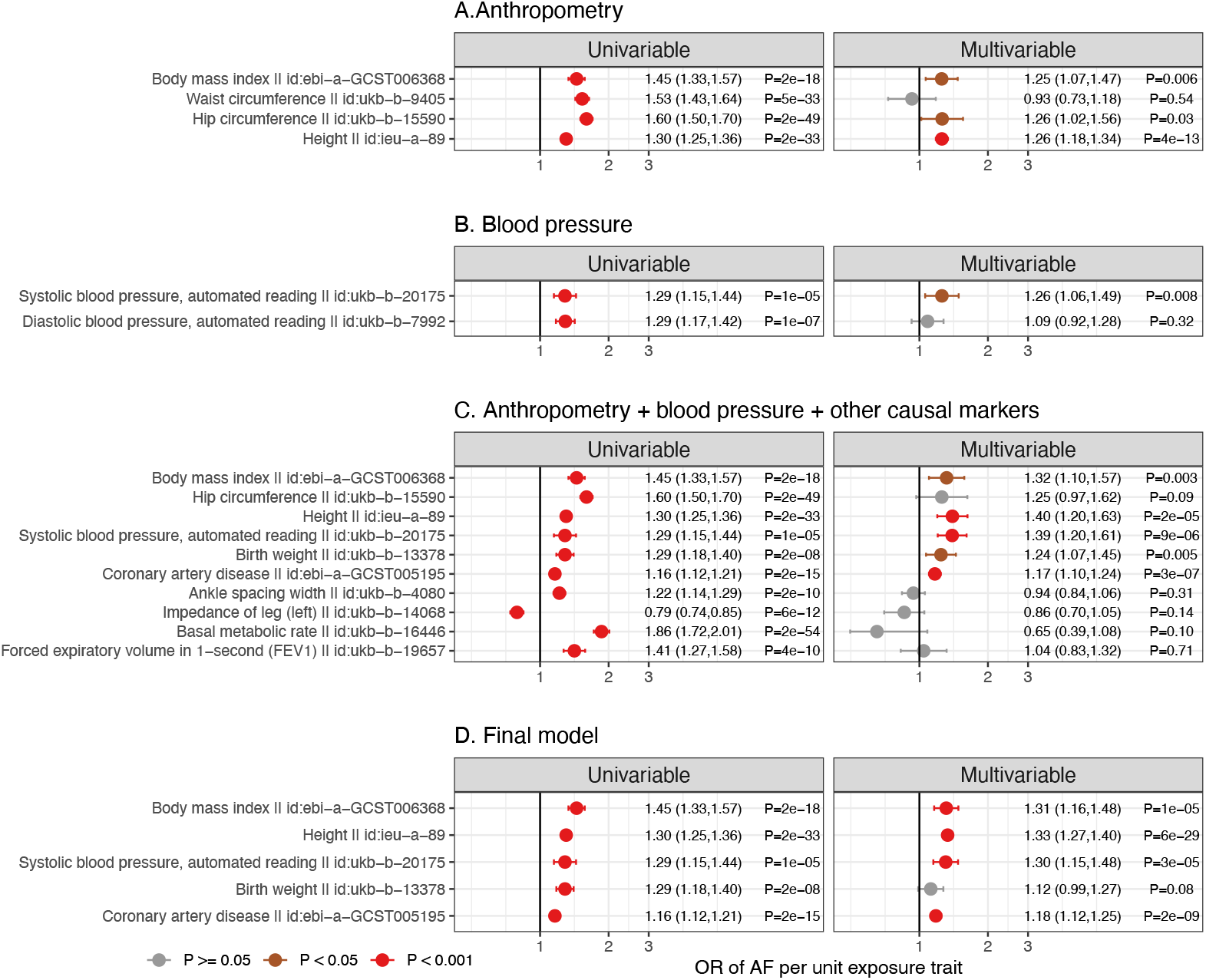
Multivariable MR analysis to identify independent causal risk factors for atrial fibrillation. In step A, we identified the independent causal risk factors among traits related to anthropometry. Causal estimates from univariable MR analysis (left) were compared to those from multivariable MR analysis (right). In step B, we similarly determined the independent risk factors for blood pressure traits. In step C, the independent causal risk factors from step A (BMI, hip and height) and B (systolic blood pressure) were then forwarded to combine with additional causal risk factors and multivariable analysis was repeated to determine their comparative casual effects. In the final step (D), independent risk factors from step C were fitted in a multivariable model. Associations with P ≥ 0.05 were coloured in grey, P < 0.05 in brown and P < 0.001 in red.

In addition, among the 2002 proteins, 578 proteins had (Steiger filtered) -*cis* instruments available and 423 of the 578 proteins had a single variant instrument. Here, MR analysis suggested 5 proteins might have causal role in AF, including higher levels of dual specificity protein phosphatase 13 isoform A (DUSP13), peptidyl−prolyl cis−trans isomerase FKBP7 (FKBP7) and spondin−1 (SPON1), and lower levels of interleukin 6 receptor (IL-6R) and tumor necrosis factor ligand superfamily member 12 (TNFSF12) (**Figure 5A**). Colocalization analysis of circulating protein levels and AF at the protein coding region further suggested that DUSP13, SPON1 and TNFSF12 had strong evidence of sharing a common causal variant (posterior probability (PP) of H4 ≥ 98%), while there was moderate evidence for IL-6R (PP of H4 = 59%) (**Figure 5B, Figure S3** and **Table S4**). However, no colocalization evidence was observed for FKBP7 (PP of H4 = 0%). To characterise the therapeutic potential of modifying these proteins in preventing AF, we looked up the drug prioritizing scores in Open Targets platform (https://www.targetvalidation.org). The results (**Figure 5C** & **Figure S4**) suggested that three out of the four proteins (all other than spondin-1) had a moderate to strong drug prioritising score for AF, ranging from 0.26 to 0.88. In particular, despite only moderate evidence of colocalization, IL-6R had the highest prioritising score among these proteins of 0.88.

**Figure 5.**
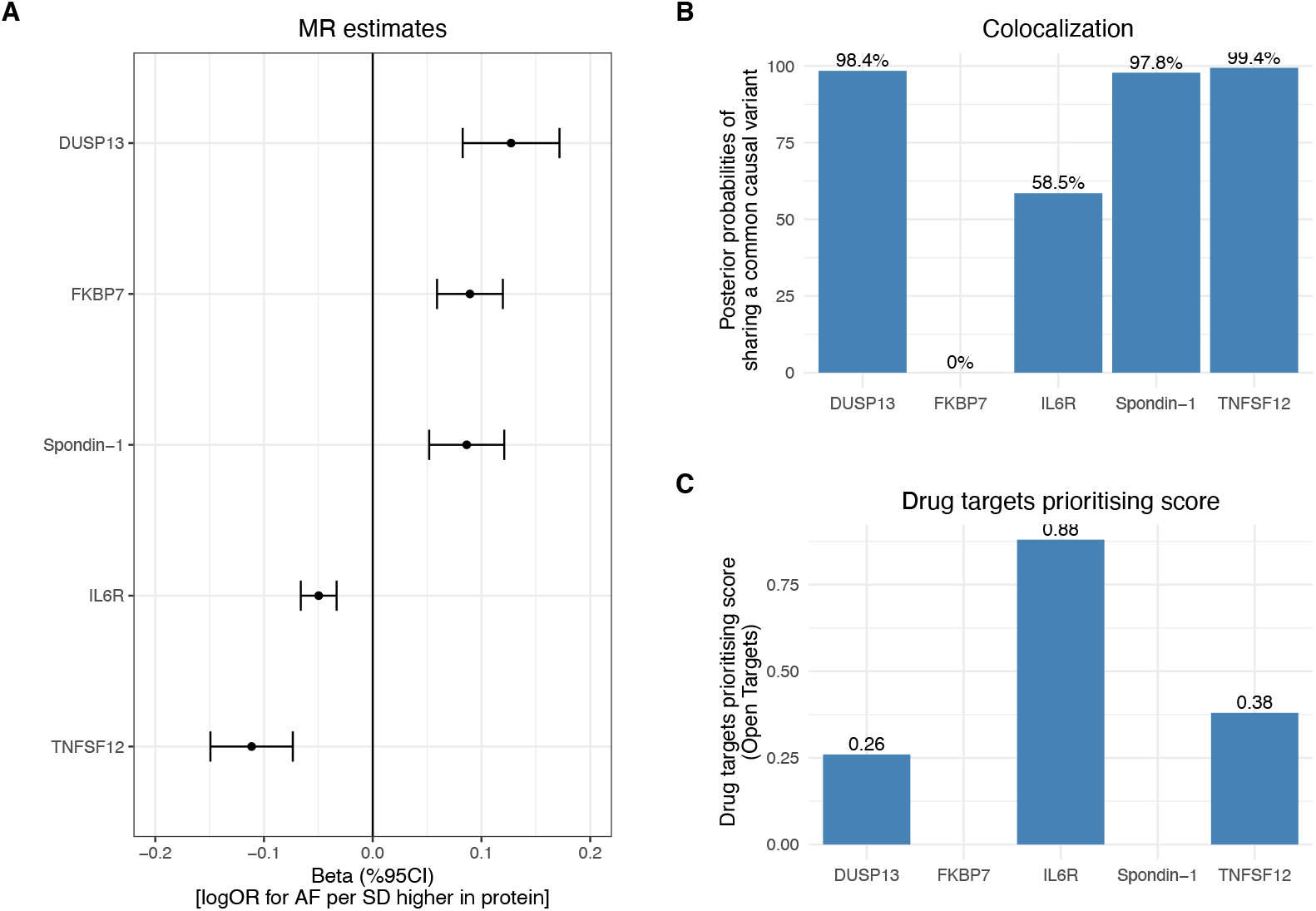
Causal role of proteins in atrial fibrillation. **A**. MR estimates of the causal effects of the proteins to AF. **B**. Colocalization analysis demonstrating the posterior probability of circulating protein and AF sharing a common causal variant at the protein coding region. Details were shown in Table S4. **C**. Drug targets prioritising score for the proteins. Data were obtained from Open Targets platform (https://www.opentargets.org). For each protein, the top 40 associated traits or diseases were illustrated in Figure S4. The platform allows prioritisation of drug targets based on the strength of their association with a disease. It allows for the prioritisation of targets by scoring target-disease associations based on evidence from 20 data sources. Similar data sources (e.g. Open Targets Genetics Portal and PheWAS) are grouped together into data types (e.g. Genetic associations). The scores for the associations range from 0 to 1; the stronger the evidence for an association, the stronger the association score (closer to 1). A score of 0 corresponds to no evidence supporting an association.

### MR analysis from atrial fibrillation to the phenome

Out of 10308 exposure-outcome pairs, 46 traits were suggested as the causal consequence of genetic liability to AF (P < 5e-6) (**Figure 6-7**). These traits include family history of heart disease or stroke, medications in relation to anticoagulation (warfarin), heart rate and blood pressure control (bisoprolol and furosemide) and antiplatelet (aspirin), and also diseases related to coronary artery disease and stroke. Results were largely consistent across the six MR methods (**Figure S5**). The causal estimates for all the 10308 exposure-outcome pairs across the six different MR methods are reported in **Table S5**.

**Figure 6.**
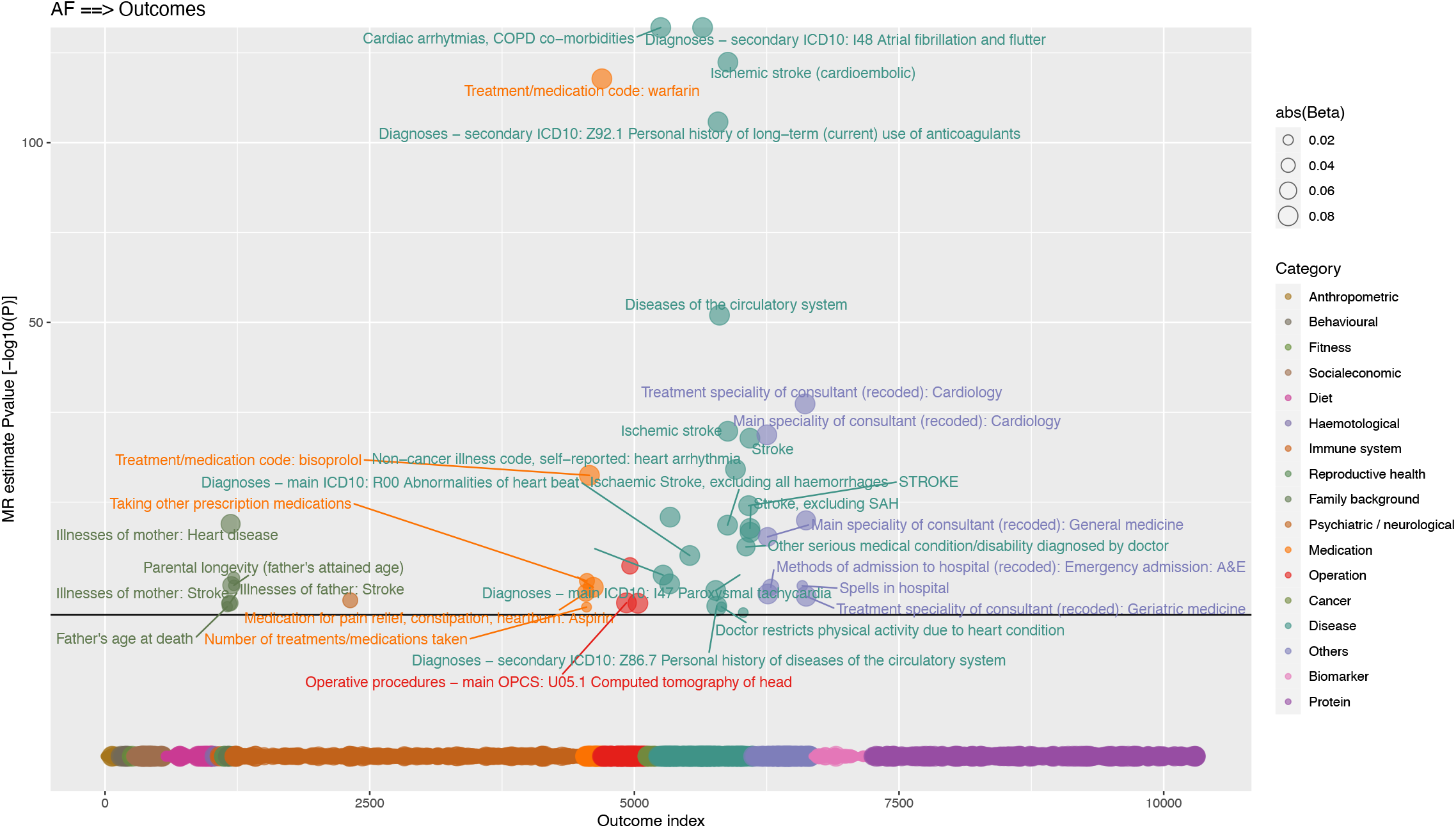
MR analysis of atrial fibrillation to phenome-wide traits. Y axis denotes the common logarithm (log10) of the P-values of the MR estimates and X axis denotes the number or index of the outcome traits. Symbols are coloured according the category of outcome traits and the symbol sizes are proportional to the absolute values of the MR estimates (based on IVW method). Horizontal black line corresponds to the Bonferroni corrected P = 0.05/10,308 traits = 5e-6. For display purposes, the associations which were not supported by any of the three-stage sensitivity test, we set the P values equal to one (indicating non-reliable).

**Figure 7.**
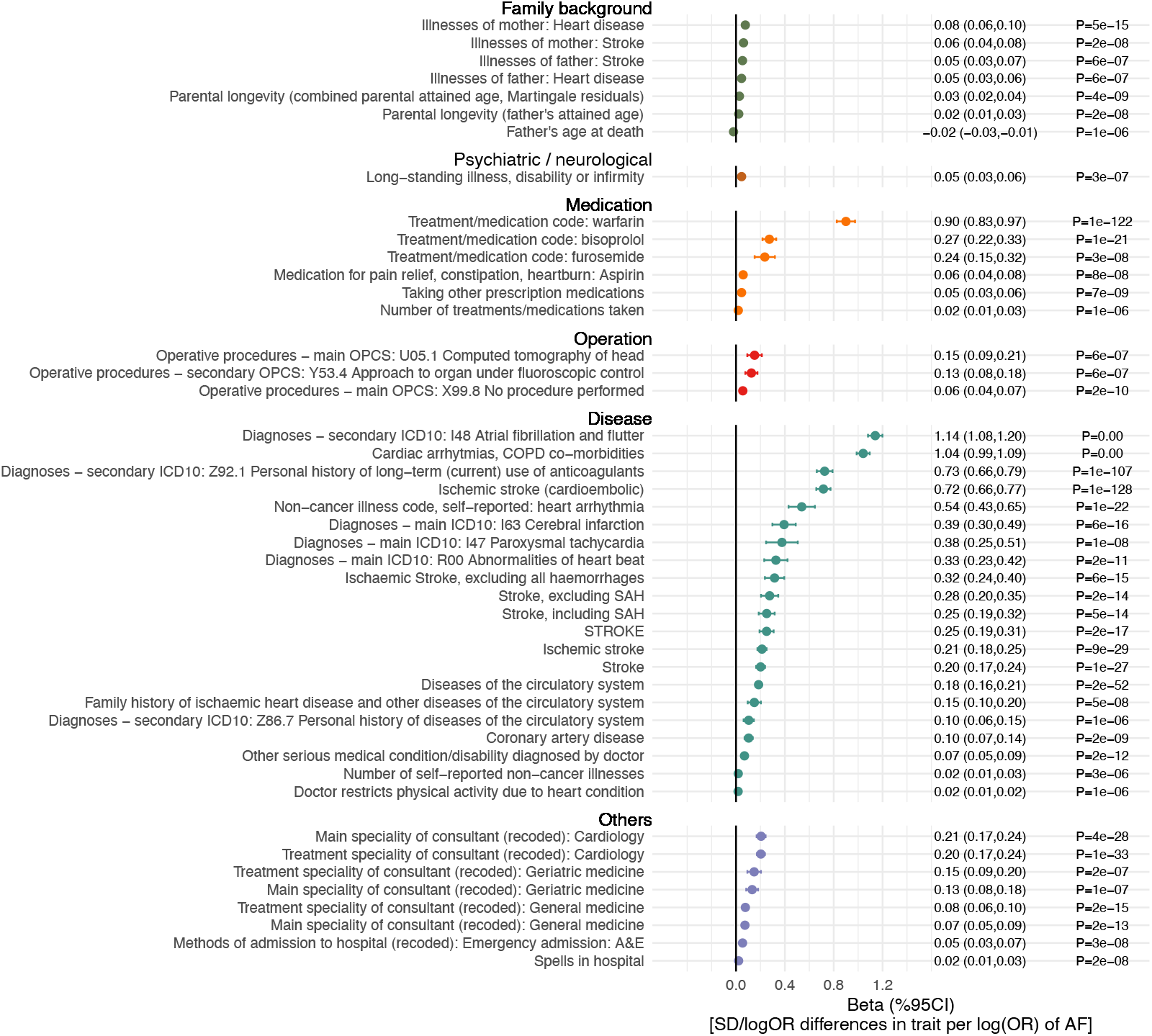
Causal consequences of genetic liability to atrial fibrillation. The associations are reported as differences in outcome traits per unit higher risk of atrial fibrillation. Traits presented are those that surpass multiple testing (Bonferroni P < 0.05/10,308 outcomes = 5e-6) in Figure 6.

As the consequences were mostly related to stroke risks or medications, we investigated whether liability to AF showed consistent association patterns across stroke types (**Figure 8**). In order to understand the degree to which genetic liability to AF contributes to stroke types and its mediating role, multivariable analyses incorporating BMI, height, SBP and CAD (independent causal risk factors for AF, Figure 4D) as the covariates were used. The conditional F-statistics for each exposure across the models are shown in **Table S6**. For stroke overall (**Figure 8A**), the results showed that genetic liability to AF retained a causal relationship in multivariable MR, and that the multivariable model further indicated that genetic liability to AF, SBP, liability to CAD each played a causal role. A similar pattern (**Figure 8B**) was also seen for ischemic stroke (the major type of stroke, accounting for around 85% of stroke cases globally).^20^ In exploring three subtypes of ischemic stroke (comprising ischemic cardioembolic ischemic stroke, large-artery atherosclerotic stroke and small-vessel stroke), different patterns (**Figure 8B1-B3**) became evident. Genetic liability to AF displayed the largest magnitude of effect with risk of cardioembolic stroke. In univariable analysis, all the risk factors displayed positive effects for cardioembolic ischaemic stroke, however in the multivariable model only genetic liability to AF retained an independent causal role (OR = 2.05 and P = 1e-89) suggesting AF might mediate the effects of the other risk factors on risk of cardioembolic ischaemic stroke (**Fig 8B1**). For large-artery atherosclerotic stroke and small-vessel ischaemic stroke, no effect of genetic liability to AF was identified in either univariable or multivariate models (**Fig 8B2** and **8B3**).

**Figure 8.**
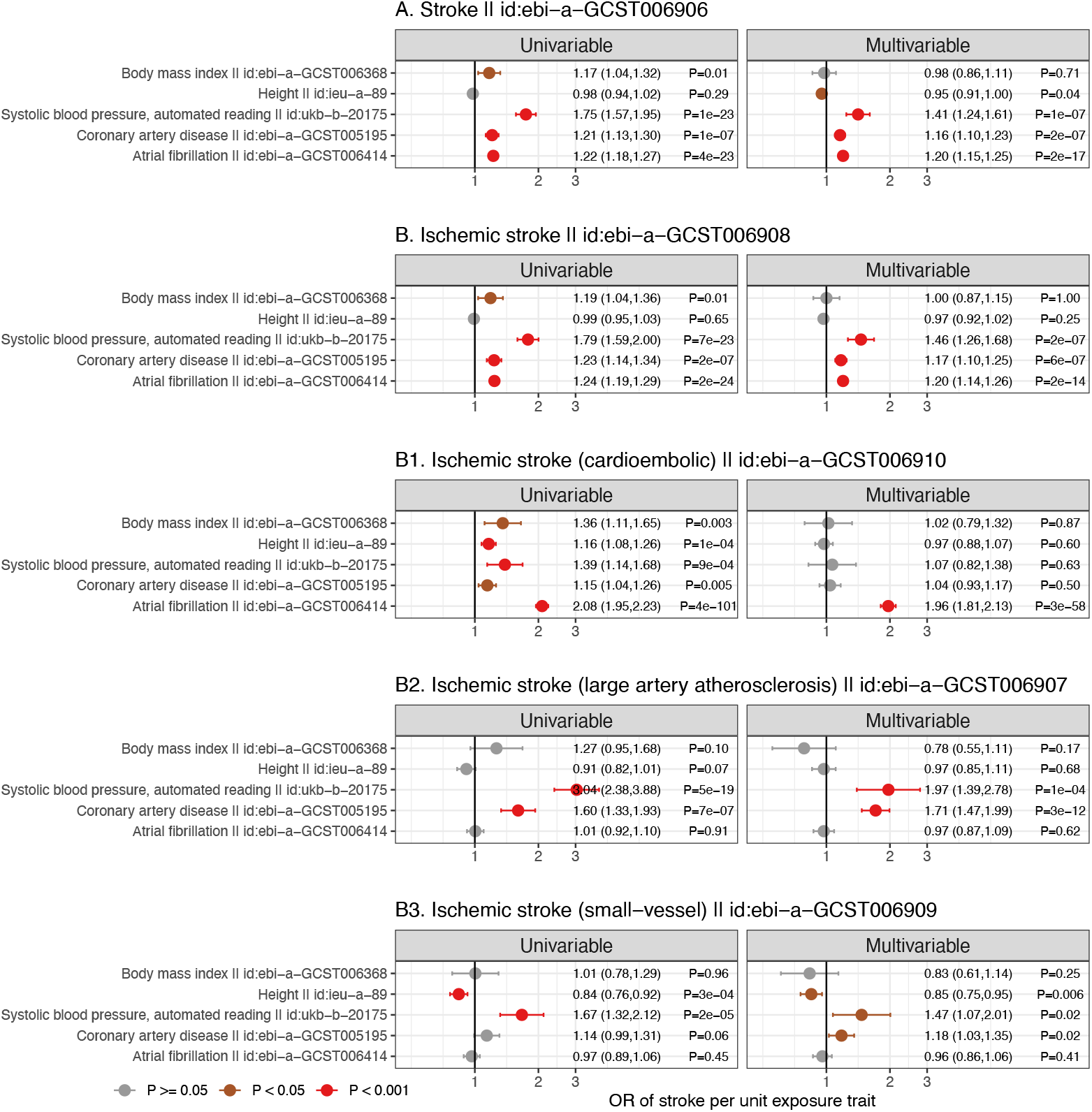
Multivariable analysis to determine independent causal risk factors for all stroke (A), ischaemic stroke (B) and ischaemic stroke subtypes (B1-B3). Causal estimates from univariable analyses (left) were compared to those from multivariable MR analyses (right). Associations with P ≥ 0.05 were coloured in grey, P < 0.05 in brown and P < 0.001 in red.

## Discussion

To systematically explore the causes and consequences of atrial fibrillation, we conducted a phenome-wide, multi-directional MR analysis of atrial fibrillation, spanning thousands of traits including anthropometric, behavioural and socioeconomic measures, diet, neurological factors, reproductive health, diseases, medication and operational codes, as well as a wide range of biomarkers and proteins. Our results suggested that adiposity indexed by BMI, height, systolic blood pressure and coronary artery disease are direct causal risk factors for risk of AF and that genetic liability to AF increases the risk of cardioembolic stroke, potentially mediating the effects of BMI, height, SBP and CAD. Several proteins, including circulating levels of IL-6 receptor, were causally related to lower risk of AF, and may have therapeutic potential.

In this work, multiple causal risk factors were identified for AF and most of them were related to adiposity. This is in line with a previous study, which found that fat and fat-free were causal for AF.^7^ In addition, our results suggest that higher height is detrimentally causal for AF,^9^ which suggests contrasting effects of height on different coronary disease: e.g. prior MR studies have shown height to be protective of coronary artery disease.^21,22^ The mechanism linking height to AF remains unclear, yet the protective effect of height for CAD has been suggested to be mediated via lower adiposity, beneficial lipid profile and better lung function.^9,21,22^ Whist our results are consistent with height not being related to all or ischaemic stroke,^22^ this overall ‘null’ effect might be driven by opposing effects on ischaemic stroke subtypes. A detailed analysis of stroke subtypes revealed distinctive patterns with higher height increasing the risk of cardioembolic stroke but lowering the risk of small vessel ischemic stroke. Our multivariable MR analysis further suggests that the positive causal role of height for cardioembolic stroke is likely to be mediated via its effect on increased risk of AF. These discrepant effects, obscured when using a composite endpoint, underscores the importance of a detailed exploration of disease subtypes.

Among the approximately 2000 proteins investigated in this study, four were suggested to be causal for AF with three (DUSP13, IL-6R and TNFSF12) displaying medium to high drug prioritising scores. Studies suggest that *DUSP13* gene expression were upregulated after stress stimulation in cardiomyocytes^23^ and that *TNFSF12* may be related to angiogenesis.^17^ Among these proteins, IL-6R showing the highest drug prioritising score of 0.88. AF have been associated with various inflammation biomarkers and a previous study implicating NLRP3 inflammasome activation (which leads to 1L-1β activation and consequently its downstream effects on IL6 acting through IL6 receptor) in AF.^24,25^ Taken together, our findings, which are consistent with a previous study^10^, underlie the therapeutic potential of pathways downstream of IL-1β for treating AF. In line with the promising genetic findings, RCTs have been conducted to assess the effects of IL-1β inhibitors in treating cardiovascular disease. In a recent phase III clinical trial, canakinumab, a monoclonal antibody inhibitor of interleukin-1 beta (IL1b), which has a license for rheumatologic disorders, was shown to lower the risk of cardiovascular diseases.^26^ Also, a recent small pilot RCT (N = 24) of canakinumab in patients with persistent AF found a numerically lower incidence of AF recurrence at 6 months in the treatment arm as compared to placebo.^27^ These initial pilot data support potential future larger trials assessing the clinical feasibility of IL-1β inhibitors (and indeed IL6R inhibition) for treating AF.

Our MR analysis of genetic liability to AF on phenome-wide traits revealed that liability to AF leads to an increased risk of stroke and stroke medications. Our results of stroke subtypes further revealed that genetic liability to AF is specifically contributing to cardioembolic ischemic stroke but not other ischaemic stroke subtypes, and it is this relationship that likely underlies the relationship of AF with combined ischaemic stroke. In contrast, a distinctive causal pattern was observed for the other two subtypes of ischemic stroke, namely large artery atherosclerosis and small-vessel disease, which are primarily dominated by blood pressure and the onset of coronary artery disease,^28^ independent of AF.

### Study limitation

The strength of this study lies in the hypothesis-free approach in assessing the multi-directional causal role of phenome-wide traits with AF, permitting the comprehensive evaluation and discoveries that we report. To ensure the robustness of the results, a three-stage sensitivity approach was designed for non-protein exposures, whist only cis acting genetic variants were used to instrument protein exposures. To address multiple testing, we used Bonferroni corrections, and the consistency of the findings was compared across 6 different MR methods. We acknowledge that sample overlapping between exposure and outcome GWAS may induce overfitting in the case of weak instrument, ^29^ however this potential bias should be marginal given the adequate F-statistics of the identified causal risk factors in the univariable MR model (**Table S3 & S6**). In addition, we used conditional F-statistics to guide our MVMR analysis in minimising bias from weak instruments. Almost all exposures had conditional F-statistics ≥ 10 except the instrument for genetic liability to CAD which had a conditional F-statistic of 9 (**Table S3 & S6**), and which showed similar estimates on univariable and multivariable MR analyses, arguing against potential weak instrument bias. Overall, despite over 5000 traits being used as exposures and over 10,000 traits being used as the outcome traits in this multi-directional MR analysis, we acknowledge that further positive findings may be revealed when more GWAS of detailed phenotypes and larger sample sizes become available.

## Conclusions

In this multi-directional MR analysis we identified adiposity, height, systolic blood pressure, and liability to coronary artery disease as independent causal risk factors for AF. Genetic liability to AF predisposes to risk of cardioembolic ischemic stroke. Several proteins, including IL-6R, may represent therapeutic potential for preventing AF, highlighting inflammation as a potential causal pathway in the aetiology of AF.

## Data Availability

All results generated from this work can be accessed in the supplement.

