## Supplementary Figures for "A phenome-wide multi-directional Mendelian randomization analysis of atrial fibrillation"

### Supplemental materials

### Content

### Supplementary tables

**(All tables are presented in excel file)**

Table S1. Causal estimates from exposures to atrial fibrillation.
Table S2. Causal estimates from typical cardiovascular risk factors to atrial fibrillation.
Table S3. F-stats for exposures in univariable and multivariable MR analysis to atrial fibrillation.
Table S4. Colocalization analysis for proteins and atrial fibrillation.
Table S5. Causal estimates from atrial fibrillation to outcomes.
Table S6. F-stats for exposures in univariable and multivariable MR analysis to stroke and stroke subtypes.

### Supplementary figures

Figure S1. Sensitivity test for the significant findings identified in the MR analysis of phenome-wide traits to atrial fibrillation.
Figure S2. Genetic correlations between the causal risk factors identified for atrial fibrillation.
Figure S3. Locuszoom plots for the proteins and Atrial Fibrillation.
Figure S4. Drug targets prioritising score for its associated traits and diseases.
Figure S5. Sensitivity test for the significant findings identified in the MR analysis of atrial fibrillation to phenome-wide traits.

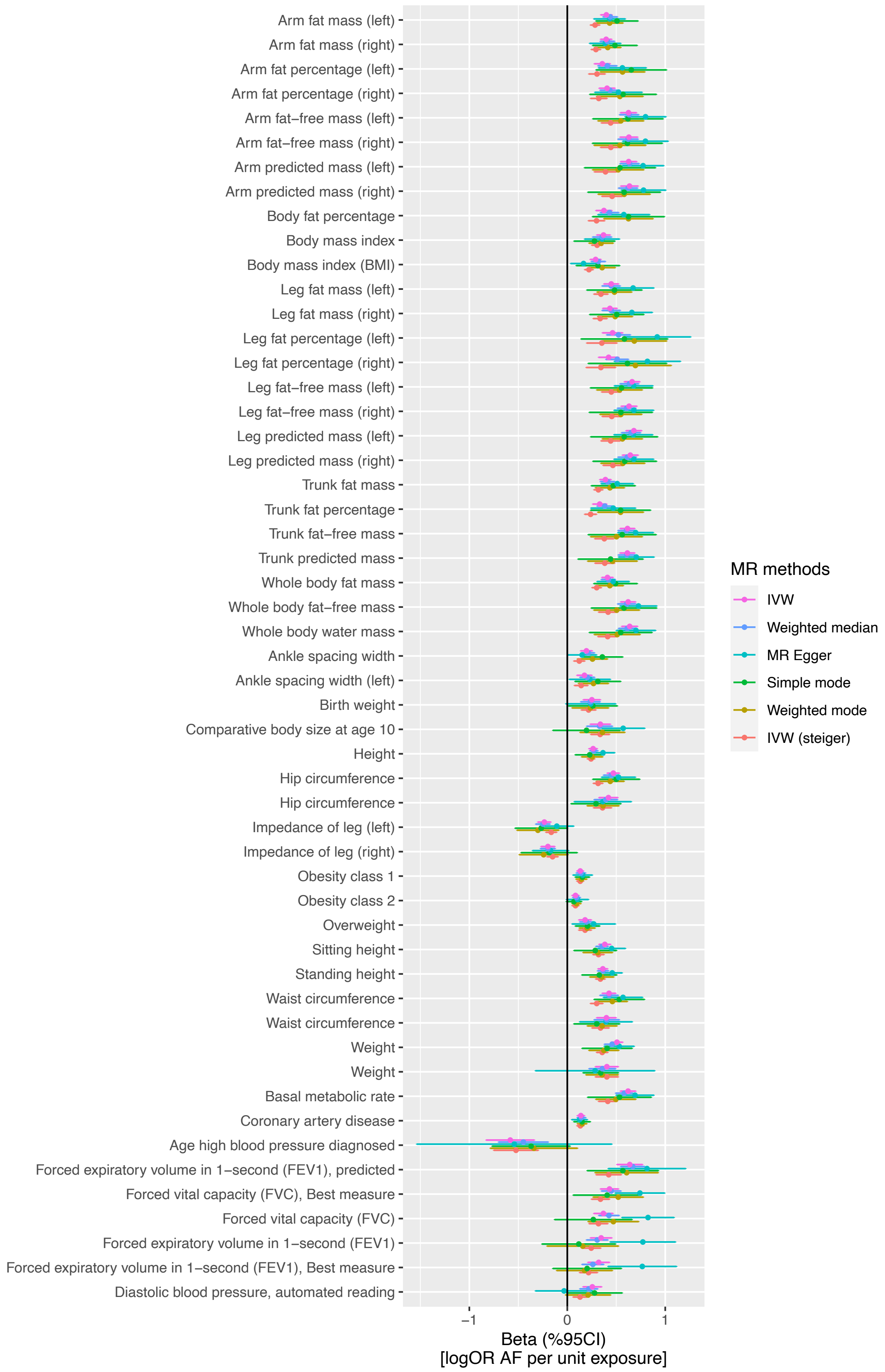


**Figure S1. Sensitivity test for the significant findings identified in the MR analysis of phenome-wide traits to atrial fibrillation.** Five different MR methods were used, including: inverse-variance weighted (IVW), MR median, MR Egger, simple mode, weighted mode and IVW with steiger filtering.

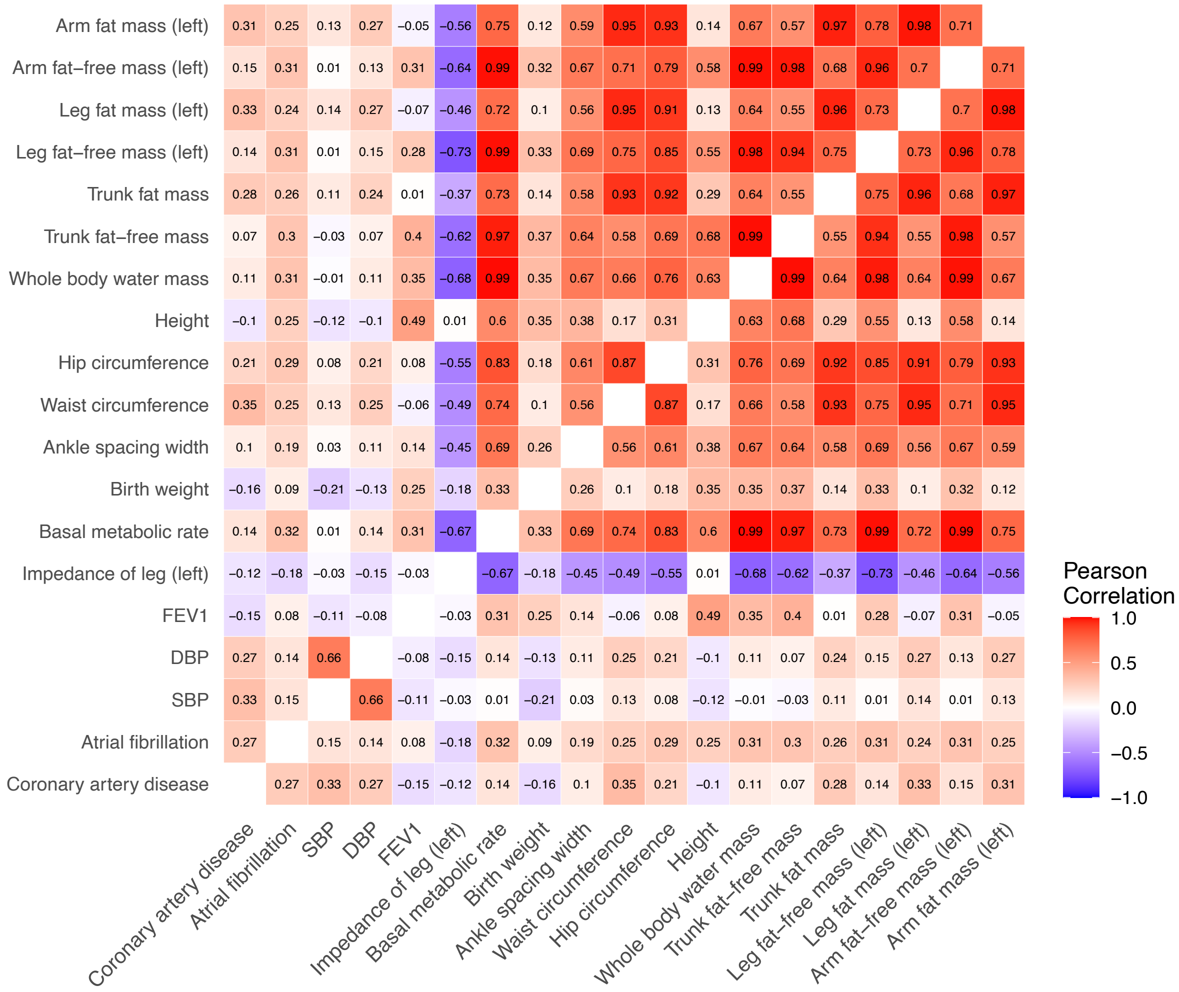


**Figure S2. Genetic correlations between the causal risk factors identified for atrial fibrillation.**

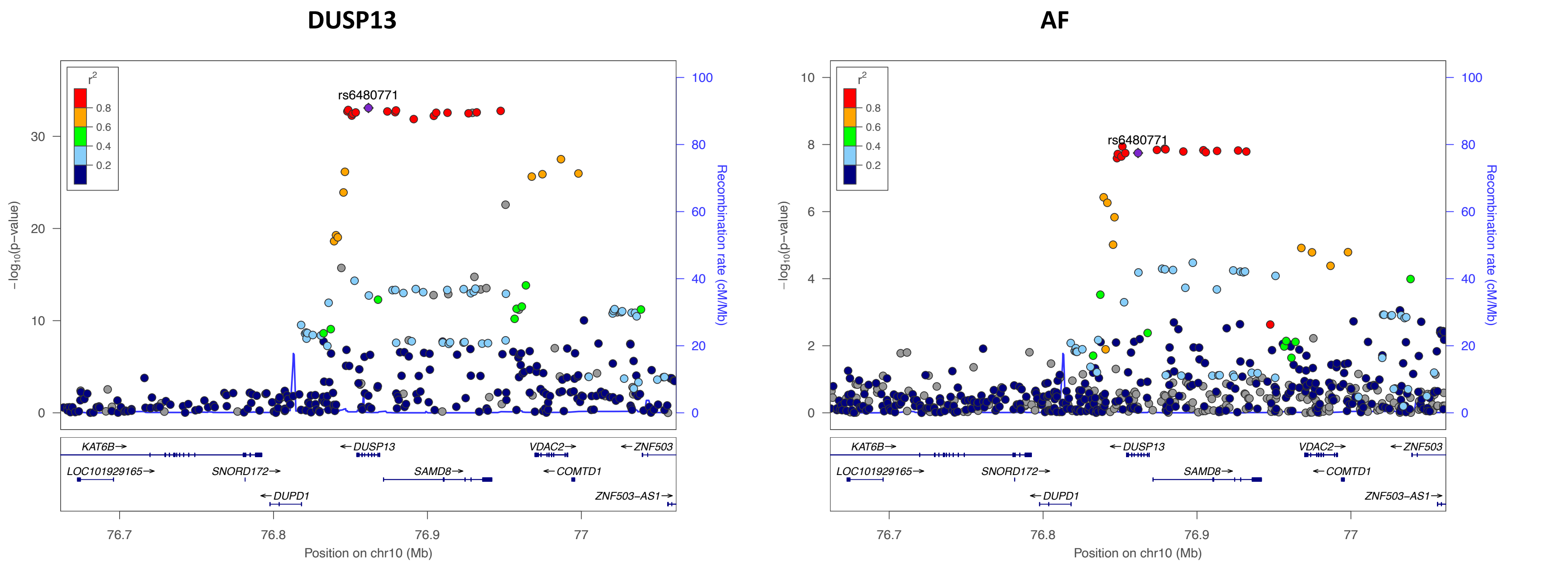


**Figure S3A**

**Figure S3A-E. Locuszoom plots for the proteins and Atrial Fibrillation.** For each protein, SNPs, 200kb-flanking of the *cis*-pQTL (purple diamond) at the coding gene were used. Corresponding region for AF were also shown.

### FKBP7

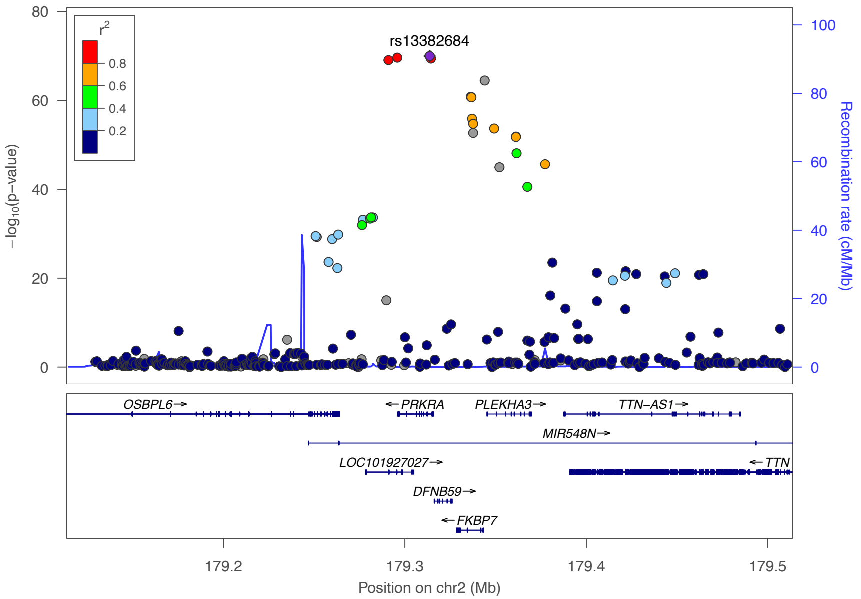


## AF

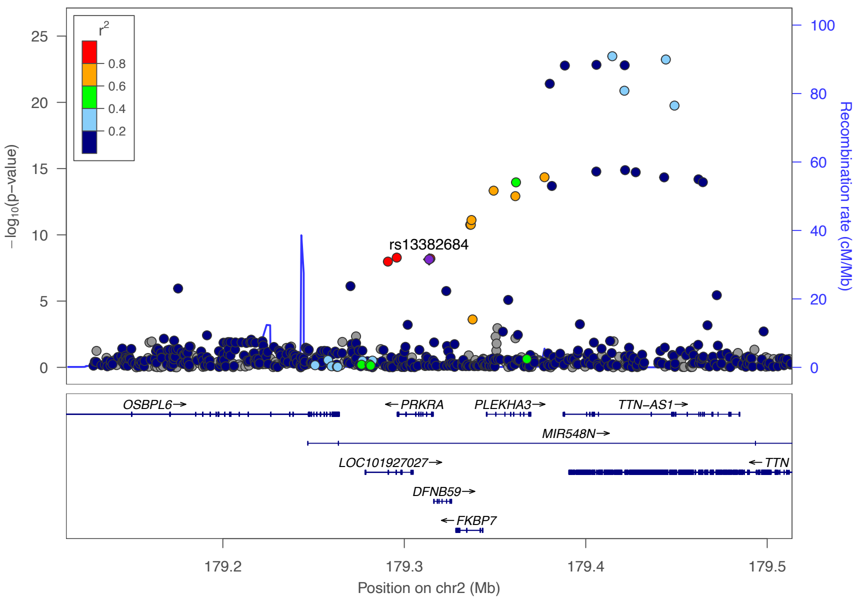


**Figure S3B**

## IL-6R

## AF

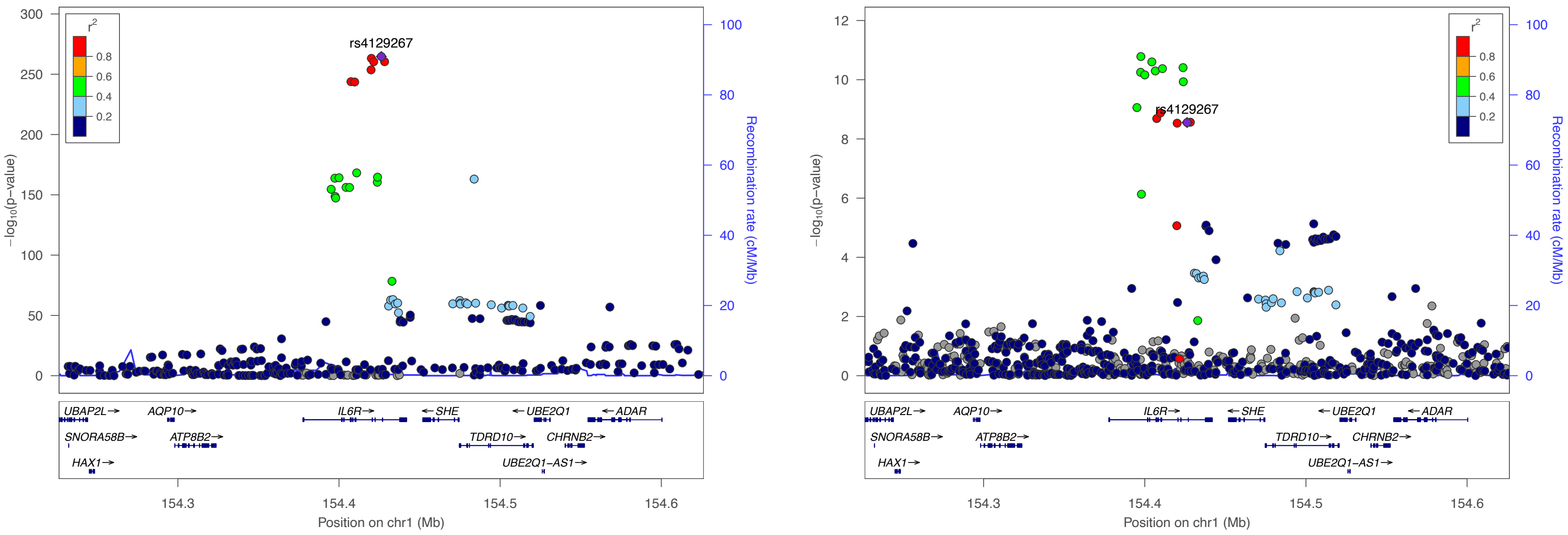


**Figure S3C**

### SPON1

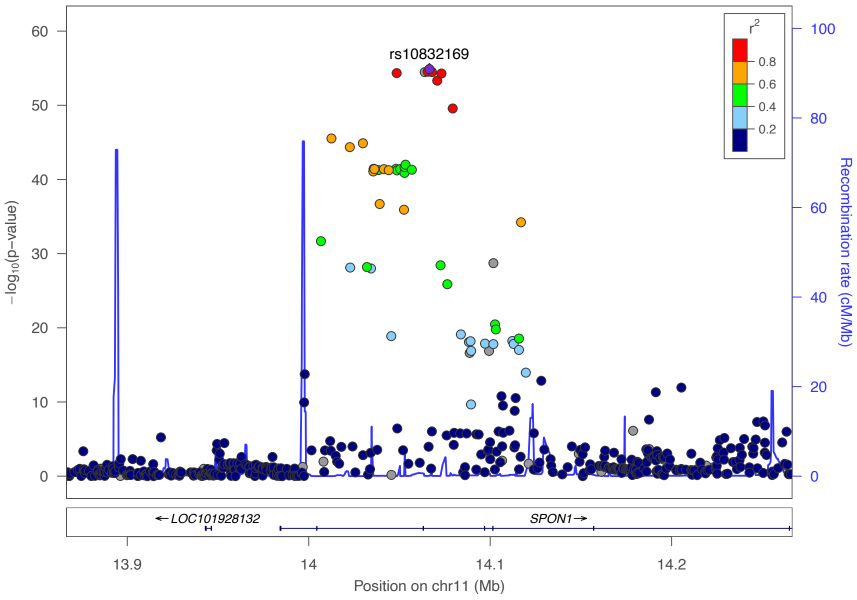


## AF

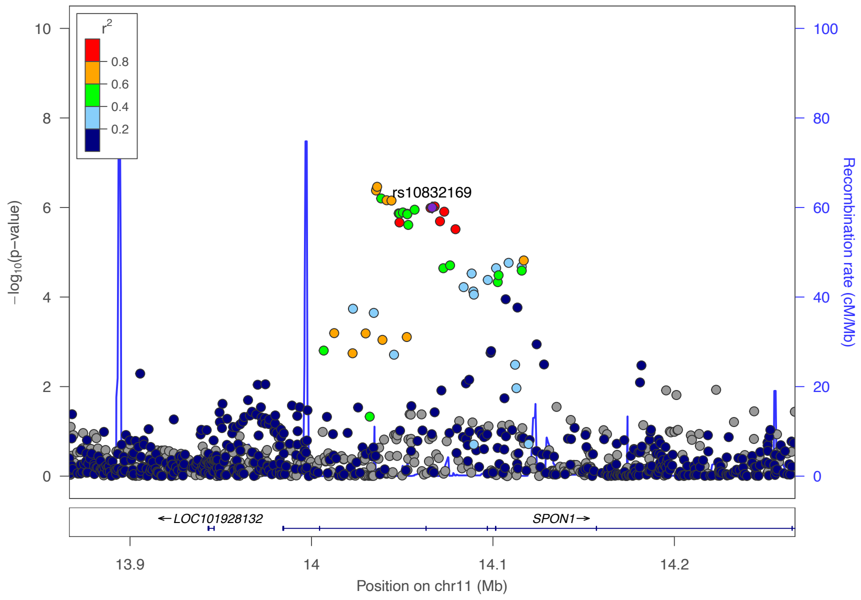


**Figure S3D**

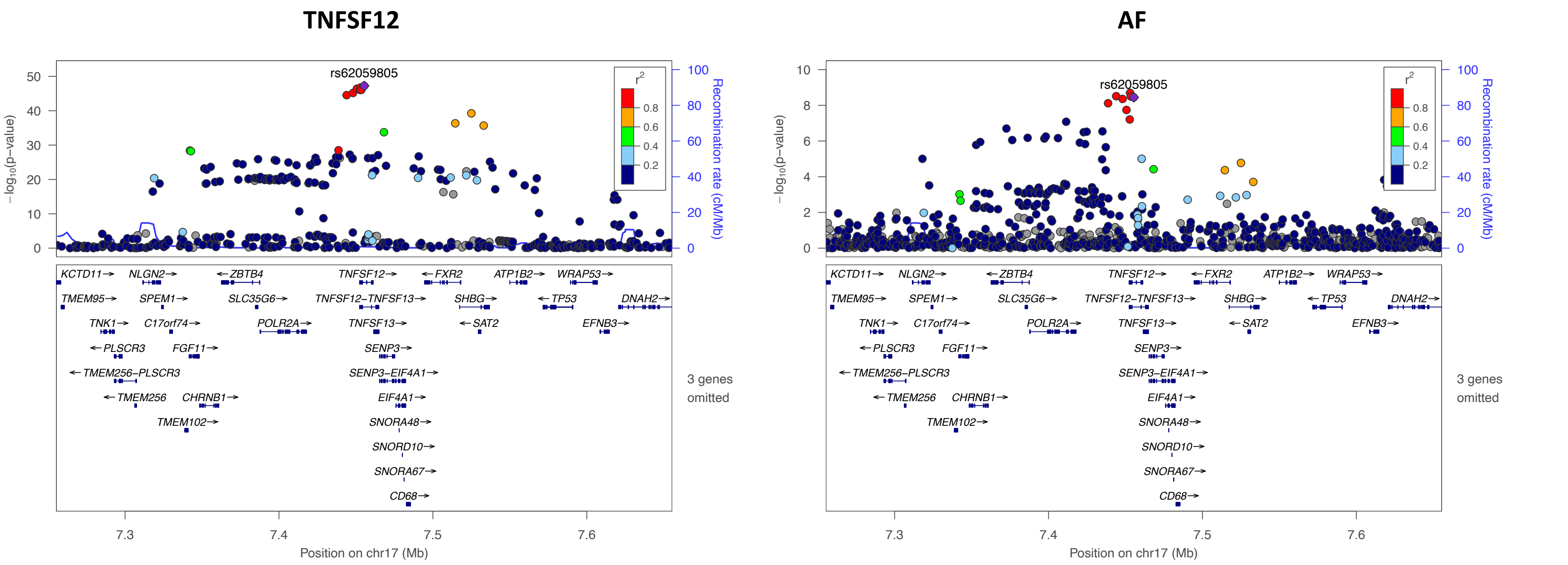
TNFSF12
AF
rs62059805
rs62059805
−log10(p−value)
Recombination rate (cM/Mb)
r2
0.8
0.6
0.4
0.2
KCTD11→ NLGN2→ ←ZBTB4 TNFSF12→ ←FXR2 ATP1B2→ WRAP53→
TMEM95→ SPEM1→ SLC35G6→ TNFSF12−TNFSF13→ SHBG→ ←TP53 DNAH2→
TNK1→ C17orf74→ POLR2A→ TNFSF13→ ←SAT2 EFNB3→
←PLSCR3 FGF11→ SENP3→
←TMEM256−PLSCR3 SENP3−EIF4A1→
←TMEM256 CHRNB1→ EIF4A1→
TMEM102→ SNORA48→
SNORD10→
SNORA67→
CD68→
3 genes omitted
7.3
7.4
7.5
7.6
Position on chr17 (Mb)

**Figure S3E**

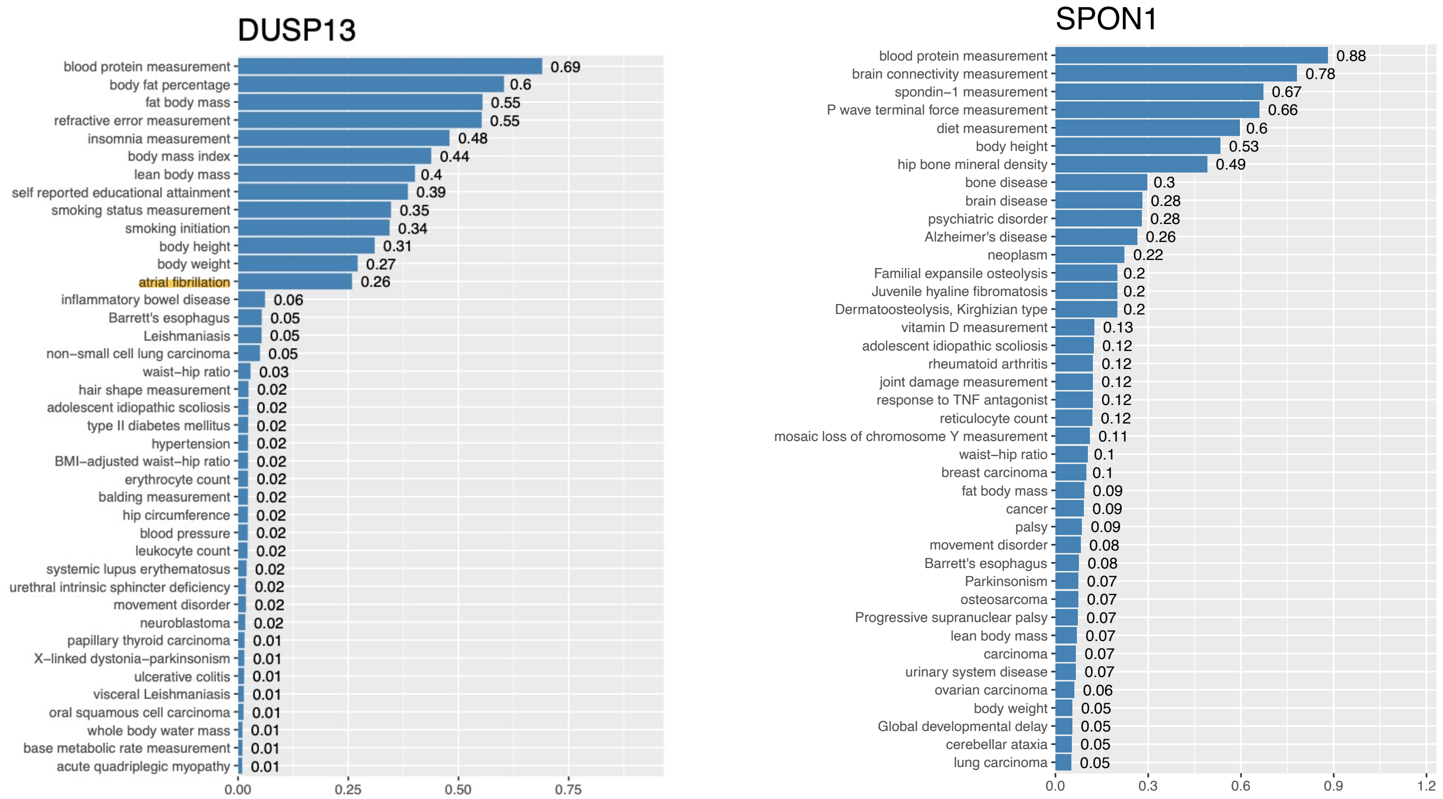


**FigureS4A**

**Figure S4A-B. Drug targets prioritising score for its associated traits and diseases.** Data were obtained from Open Targets platform (https://www.opentargets.org). For each protein, the top 40 associated traits or diseases were illustrated here. The platform allows prioritisation of drug targets based on the strength of their association with a disease. It allows for the prioritisation of targets by scoring target-disease associations based on evidence from 20 data sources. Similar data sources (e.g. Open Targets Genetics Portal and PheWAS) are grouped together into data types (e.g. Genetic associations). The score for the associations ranges from 0 to 1; the stronger the evidence for an association, the stronger the association score (closer to 1). A score of 0 corresponds to no evidence supporting an association.

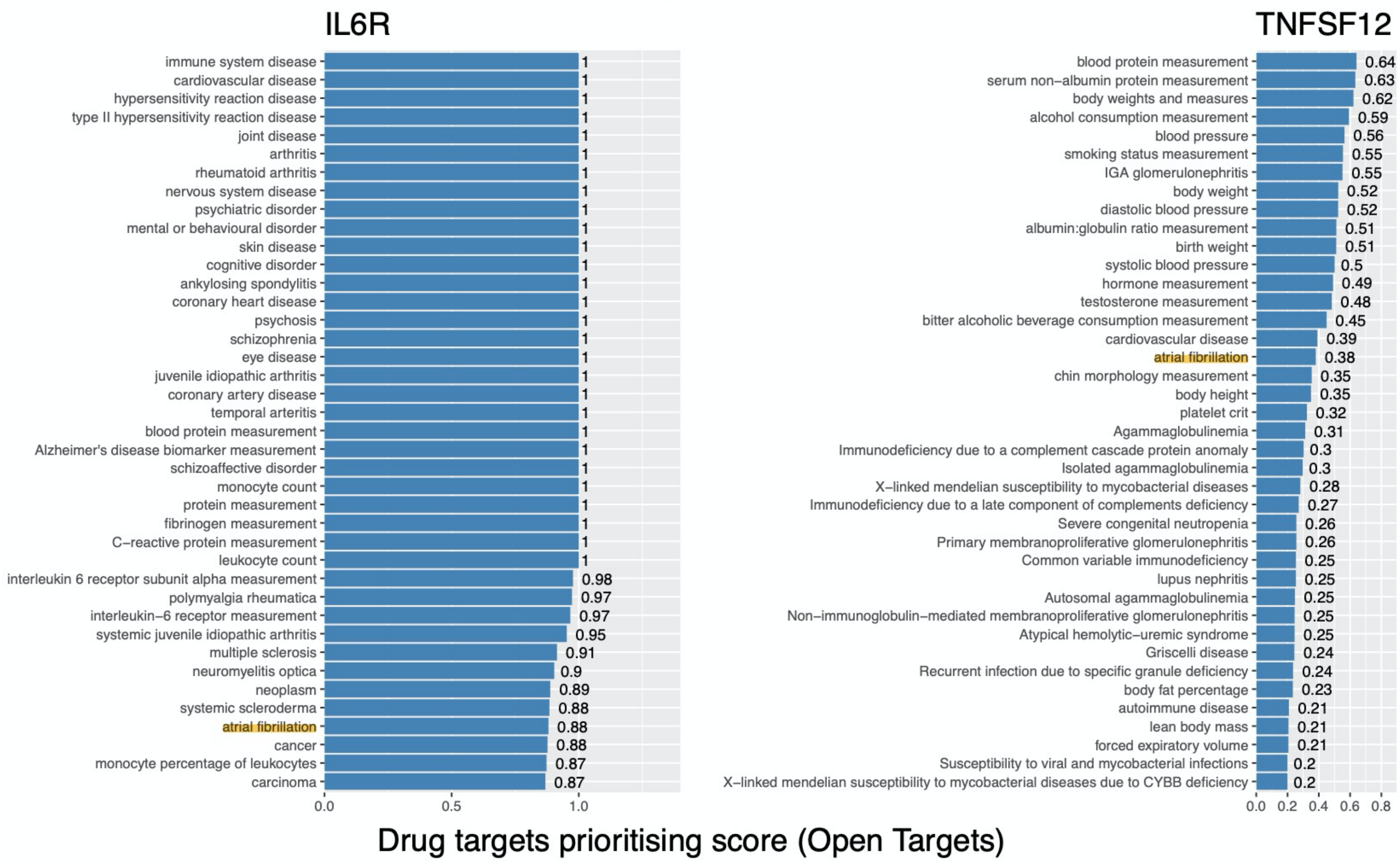

IL6R
immune system disease 1
cardiovascular disease 1
hypersensitivity reaction disease 1
type II hypersensitivity reaction disease 1
joint disease 1
arthritis 1
rheumatoid arthritis 1
nervous system disease 1
psychiatric disorder 1
mental or behavioural disorder 1
skin disease 1
cognitive disorder 1
ankylosing spondylitis 1
coronary heart disease 1
psychosis 1
schizophrenia 1
eye disease 1
juvenile idiopathic arthritis 1
coronary artery disease 1
temporal arteritis 1
blood protein measurement 1
Alzheimer's disease biomarker measurement 1
schizoaffective disorder 1
monocyte count 1
protein measurement 1
fibrinogen measurement 1
C−reactive protein measurement 1
leukocyte count 1
interleukin 6 receptor subunit alpha measurement 0.98
polymyalgia rheumatica 0.97
interleukin−6 receptor measurement 0.97
systemic juvenile idiopathic arthritis 0.95
multiple sclerosis 0.91
neuromyelitis optica 0.9
neoplasm 0.89
systemic scleroderma 0.88
atrial fibrillation 0.88
cancer 0.88
monocyte percentage of leukocytes 0.87
carcinoma 0.87
0.0 0.5 1.0
TNFSF12
blood protein measurement 0.64
serum non−albumin protein measurement 0.63
body weights and measures 0.62
alcohol consumption measurement 0.59
blood pressure 0.56
smoking status measurement 0.55
IGA glomerulonephritis 0.55
body weight 0.52
diastolic blood pressure 0.52
albumin:globulin ratio measurement 0.51
birth weight 0.51
systolic blood pressure 0.5
hormone measurement 0.49
testosterone measurement 0.48
bitter alcoholic beverage consumption measurement 0.45
cardiovascular disease 0.39
atrial fibrillation 0.38
chin morphology measurement 0.35
body height 0.35
platelet crit 0.32
Agammaglobulinemia 0.31
Immunodeficiency due to a complement cascade protein anomaly 0.3
Isolated agammaglobulinemia 0.3
X−linked mendelian susceptibility to mycobacterial diseases 0.28
Immunodeficiency due to a late component of complements deficiency 0.27
Severe congenital neutropenia 0.26
Primary membranoproliferative glomerulonephritis 0.26
Common variable immunodeficiency 0.25
lupus nephritis 0.25
Autosomal agammaglobulinemia 0.25
Non−immunoglobulin−mediated membranoproliferative glomerulonephritis 0.25
Atypical hemolytic−uremic syndrome 0.25
Griscelli disease 0.24
Recurrent infection due to specific granule deficiency 0.24
body fat percentage 0.23
autoimmune disease 0.21
lean body mass 0.21
forced expiratory volume 0.21
Susceptibility to viral and mycobacterial infections 0.2
X−linked mendelian susceptibility to mycobacterial diseases due to CYBB deficiency 0.2
0.0 0.2 0.4 0.6 0.8
Drug targets prioritising score (Open Targets)


**Figure S4B**

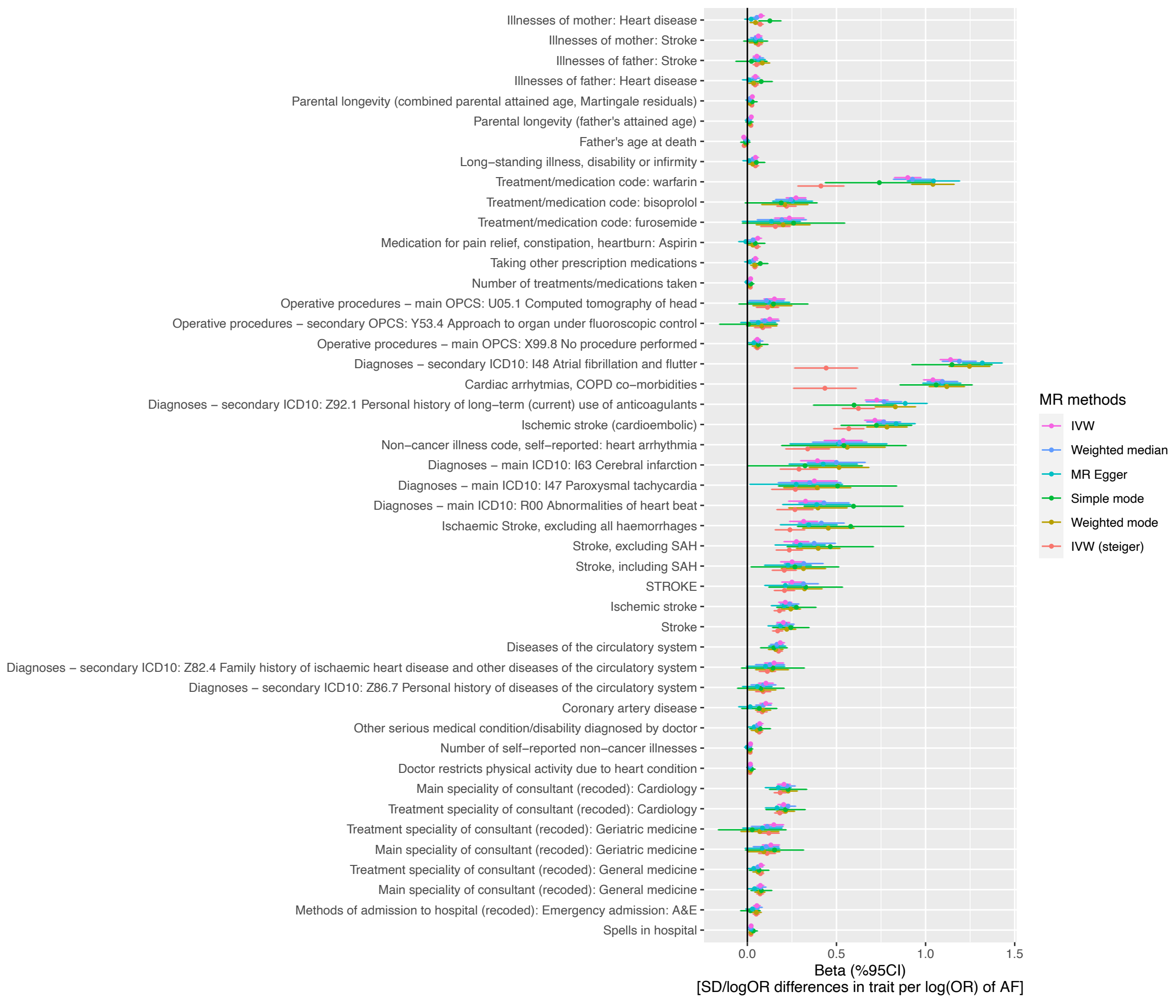


**Figure S5. Sensitivity test for the significant findings identified in the MR analysis of atrial fibrillation to phenome-wide traits.** Five different MR methods were used, including: inverse-variance weighted (IVW), MR median, MR Egger, simple mode, weighted mode and IVW with steiger filtering.
